# Knowledge, attitude and practice towards antibiotic use and resistance among the veterinarians in Bangladesh

**DOI:** 10.1101/2021.06.03.21258263

**Authors:** Md Samun Sarker, Iftekhar Ahmed, Nure Alam Siddiky, Shariful Islam, Ruhena Begum, Ayesha Ahmed, Fatema Akter Mahua, Md Ehsanul Kabir, Mohammed A. Samad

**Affiliations:** Antimicrobial Resistance Action Center (ARAC), Bangladesh Livestock Research Institute (BLRI), Savar, Dhaka-1341, Bangladesh; Department of Pharmacy, Jahangirnagar University, Bangladesh; Institute of Epidemiology, Disease Control and Research (IEDCR), Mohakhali-1212, Bangladesh; Department of Public health and Informatics, Jahangirnagar University, Bangladesh

## Abstract

**Background:** The emergence of antimicrobial resistance is growing human and animal health concern around the world. When a number of studies have emphasized the Knowledge, Attitude and Practice (KAP) regarding antibiotic use and resistance in humans, little attention has been paid to the veterinary sector. The aim of this study was to understand the KAP towards antibiotic use and resistance among the veterinarians in Bangladesh.

**Methods:** A cross-sectional online based questionnaire survey was conducted from August to September 2020 among the registered veterinary practitioners. A self-administered Google form questionnaire consists of 45 questions on knowledge, attitude and practice regarding antibiotic use and resistance were used.

**Results:** A total of 208 registered veterinarians participated in this study. 85.1% of the participants were male and 54.8% of the participants had a Masters degree. Around 52% of the veterinarians were poultry practitioners. All respondents were familiar with antimicrobials. The participants (91.4%) knew that antibiotics can not cure viral infections while 97.6% believed that frequent antibiotic prescription rendered them less effective. Participants claimed that only they are eligible to prescribe drugs for the treatment of animals and around 80% disagreed with adding antibiotics with feed/water as a growth promoter in livestock. Of the total participants, 87% believed that a local antimicrobial guideline would be more effective than an international one. However, gaps in practices were highlighted in our study, suggesting training deficiencies.

**Conclusion:** The present study for the first time conducted in Bangladesh dictates the future interventions like academic courses, workshops, and seminars on antibiotic usage and resistance are needed to ameliorate the knowledge, behavior and practice of veterinarians with regards to the rational use of antibiotics.

## Introduction

The global development of intensive farming has led to an upsurge in antimicrobial use (AMU) that leads to the emergence and spread of antimicrobial resistance (AMR) [1]. Antibiotics are used as therapeutic as well as growth promotion purposes in animal farming practices in Bangladesh. Irrational use of antibiotics in animals is considered one of the key drivers of AMR evolution [2]. Worldwide consumption of antibiotics in animals is very high and it is expected to rise 67% by 2030 [1]. Most antibiotics are used in both human and animal interface, so the emergence of resistance through veterinary use is likely to have consequences on human health [3-5]. Bangladesh has been experiencing a high incidence of AMR [6,7]. Misuse and abuse of antibiotics are common both in humans and in animals in Bangladesh [6, 8-10]. A study with 73 poultry farms in Bangladesh reported usage of antibiotics without prescribing by registered veterinarians [10]. The same study found the presence of residual antibiotics in 26% of the tested samples (breast, thigh and liver) [10]. A study revealed that majority of the antibiotics used in the poultry farms were falls under Watch and Reserve group rather than Access [11]. Antibiotics in the Watch group have a greater resistance potential and comprise the majority of the top priority agents among the critically important antimicrobials for human medicine and/or antibiotics that are at reasonably high risk of bacterial resistance selection. Reserve group antibiotics should be reserved for the treatment of infections caused by multidrug-resistant organisms, whether confirmed or suspected. Antibiotics in the reserve category should be used only as a ‘last resort’. Antibiotics in the access group are active against a broad variety of regularly encountered susceptible infections while also having a lesser resistance potential than antibiotics in the other categories. The findings of other studies identified that registered veterinarians also prescribed higher classes of antibiotics widely for treating healthy and unhealthy poultry flocks [12,13].

The WHO recommends an overall reduction of medically important antimicrobial use in food-producing animals, as well as a complete cessation for disease prevention and growth promotion of food-producing animals [14]. The Government of Bangladesh has enacted “animal and fish feed act 2010” which prohibited the use of all antibiotics in animal and fish feed [15], but subsequent studies showed that such laws were not properly enforced and the use of antibiotics with animal feeds is quite common [8–10]. This shows that unless raising awareness, motivational and ownership among the veterinarians, farmers, feed sellers, drug sellers, misuse of antibiotics in livestock will most likely continue to persist.

Any change in practice must start with the animal healthcare providers, followed by changes in the antibiotic usage among the farmers. To make effective and sustainable strategies, recommendations and treatment guidelines to maximize the therapeutic efficacy and reduce AMR in both human and animals, assessing the knowledge, attitude and practice (KAP) of veterinary practitioners are pertinent. Many countries has already been conducted similar studies to do deep this issue in a numbers perspective related to human and animals [16–20]. Nevertheless, such study is still lacking from Bangladesh. In Bangladesh, the KAP regarding antibiotic use and resistance among veterinary students has been reported previously [21]. Therefore, in this study, we have explored the KAP of the veterinarians of Bangladesh regarding antibiotic use and resistance. To our knowledge, this is the first antibiotic KAP study among the Bangladeshi veterinarians.

## Methods

### Study design and sampling

A cross-sectional study was conducted from August to September, 2020 using a self administered questionnaire. Using a purposive sampling, data collection was conducted among the registered veterinary practitioners of Bangladesh listed by the Bangladesh Veterinary Council (BVC)-government regulatory body of veterinary legislation and certification for veterinary practices in Bangladesh. To date, there are 7536 registered veterinarian in Bangladesh. BVC listed veterinarians are only certified person to treat and prescribe the antibiotics for the animals. Through social media(Facebook, LinkedIn and WhatsApp), the questionnaire was posted and circulated in different veterinary professional groups. The social media based survey was launched on August 05, 2020. In mid-September, 2020, the online questionnaire link of questionnaire was also messaged via mobile and facebook messanger text and emailed to each registered veterinarian to boost the response rate. Regarding the sample size, the aim was to include as many veterinarians as possible.

### Questionnaire development

An online based approximate 20-30 minutes long questionnaire was developed using the Google Forms platform by a multidisciplinary team consisting of microbiologist, public health specialist and epidemiologist. The questionnaire comprised of four sections: the first one consisted of the demographic information of the veterinarians, second section included 13 questions on knowledge on antibiotics and AMR, third section contained 14 questions on attitudes and fourth section had 18 questions on practices regarding antibiotic use and their resistance. The majority of the answers were in multiple choice format. The questionnaire was pretested among the veterinarians at the Antimicrobial Resistance Action Center (ARAC), Bangladesh Livestock Research Institute (BLRI) to validate and finalize the questionnaire. Finally, minor changes has been made in compliance with participants response and was circulated to the registered veterinarians. All the participants from ARAC in the pilot study were reminded to not participate in the final survey. Respondents were asked to read the consent paper and made aware of the purpose of the study and requested to provide consent before participating in the survey. The participation in the survey was completely non-compulsory and unpaid. If participants were willing to participate in the survey, they were first asked to click the “Yes” button as a consent before continuing to the next sections. To prevent the loss of confidentiality, no identifying information of the participant was collected. In addition, all data were exported into an excel file and given a serial number. This data was only accessible to the research team and investigators.

### Ethical considerations

The study protocol was reviewed and approved by the ARAC, Animal Health Research Division, BLRI, Bangladesh (Approval no: 05/06/2020:06).

### Statistical analysis

Quantitative data were entered into MS excel-2013 (Microsoft Corporation, Redmond, WA 98052, USA) and analyzed in STATA/IC-13 (StataCorp, 4905, Lakeway Drive, College station, Texas 77845, USA). Descriptive analysis was conducted to determine the frequency and percentage of responses regarding KAP. We used Chi-square test or the Fisher exact test to identify the potential association between variables with different age groups, field of expertise, type of service and years of experience of the veterinary practitioners. The statistical significance was set at *p* ≤0.05.

## Results

### Participants’ characteristics

A total number of 208 veterinarians responded and took part in the questionnaire survey from eight administrative divisions of all Bangladesh. Most of the participants were male (177/208; 85.1%), and 93.8% (195/208) were at the age from 25-35 years old. About 44% (92/208) had a Doctor of Veterinary Medicine (DVM) degree while about 55% (114/208) had a Master’s degree. Half of the veterinarians were poultry practitioners and the rest were pet animals and large and small animals practitioners. Around 31% (65/208) had an experience of greater than 5 years. Detailed characteristics of the participants are presented in Table 1.

**Table 1.** Sociodemographic characteristics of the participants.

| Characteristic | Number (n) | Percentage (%) |
| --- | --- | --- |
| Gender |  |  |
| Male | 177 | 85.1 |
| Female | 31 | 14.9 |
| Education |  |  |
| DVM | 92 | 44.2 |
| Masters | 114 | 54.8 |
| PhD | 2 | 0.96 |
| Age |  |  |
| 25-30 Years | 109 | 52.4 |
| 31-35 Years | 86 | 41.4 |
| Above 36 Years | 13 | 6.3 |
| Field of expertise |  |  |
| Poultry practitioner | 108 | 51.9 |
| Pet animals' practitioner | 18 | 8.7 |
| Large & Small Animals' Practitioner | 82 | 39.4 |
| Type of Service |  |  |
| Private | 155 | 74.5 |
| Governmental | 53 | 25.5 |
| Years of practice |  |  |
| >1 Year | 39 | 18.8 |
| 1-3 Year | 63 | 30.3 |
| >3-5 Years | 41 | 19.7 |
| Above 5 Years | 65 | 31.3 |
| Job location (Division) |  |  |
| Barisal | 8 | 3.9 |
| Chittagong | 71 | 34.1 |
| Dhaka | 59 | 28.4 |
| Khulna | 3 | 1.4 |
| Mymensingh | 15 | 7.2 |
| Rajshahi | 30 | 14.4 |
| Rangpur | 17 | 8.2 |
| Sylhet | 5 | 2.4 |

### Knowledge on antibiotics and AMR

Almost all respondents were familiar with antimicrobials and antibiotics, but 17.3% were unaware that antibiotics are different from antimicrobials. Although most participants (91.4%) knew that antibiotics cannot cure viral infections, 33.6% believed the use of antibiotics would speed up recovery from common cold, cough, and other viral infections. The vets (47.6%) who had experienced of more than 1 to 3 years of practicing thought antibiotics would be effective against common cold, cough and viral infections. All of the vets were aware of antibiotic resistance and 97.6% knew that frequent prescription of antibiotics can render them less effective. However, some practitioners (6.7%) were unaware of the concept of antibiotic susceptibility testing. In addition, all vets regardless of age, field of expertise, type of service and experience believed biosecurity and improved hygiene could reduce the use of antibiotics (Table 2).

**Table 2.**
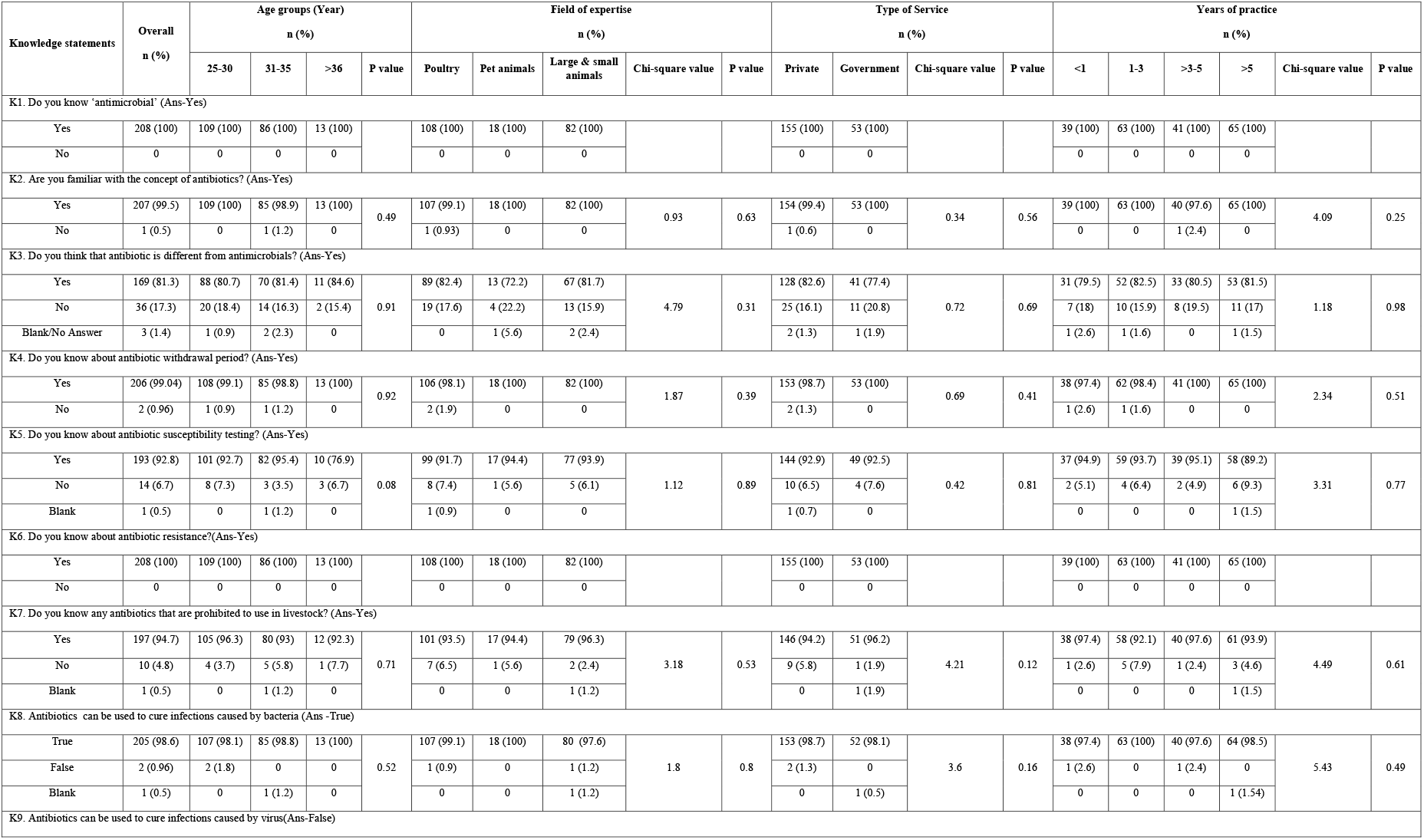
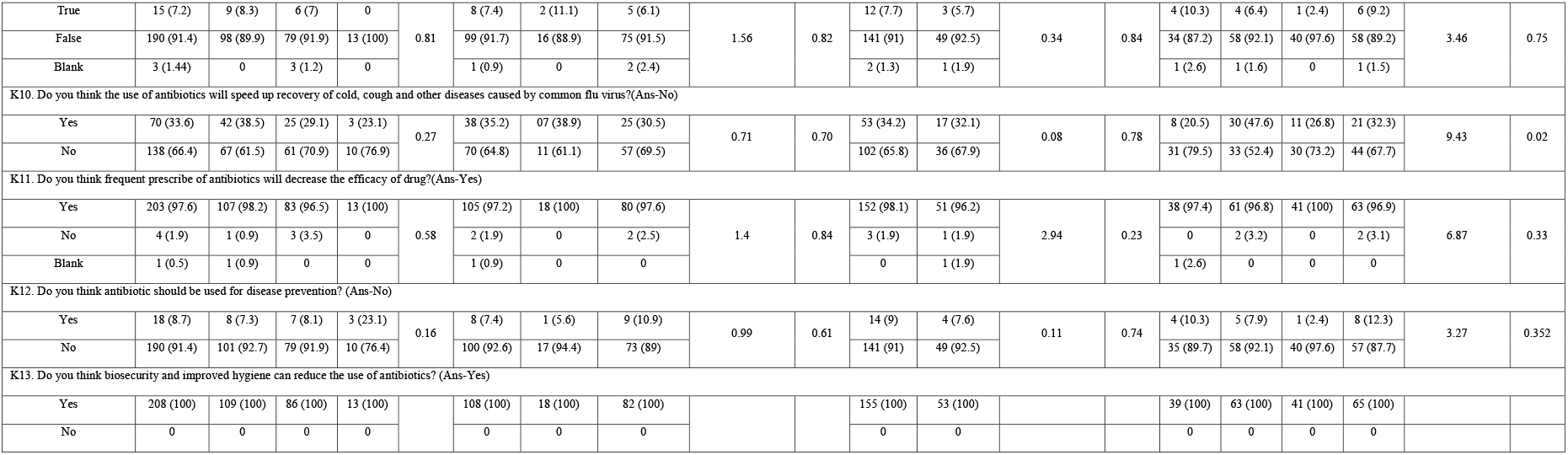
Veterinarian’s knowledge on antibiotic use and resistance.

### Attitude towards antibiotic use and resistance

Out of the 208 participants, 207 opinioned that only veterinarians are eligible to prescribe drugs for animals. Moreover, nearly all agreed that antibiotic abuse is prevalent in veterinary practices in Bangladesh. Practitioners also had a positive attitude towards vaccination for the purpose of preventing diseases and for reducing the use of antibiotics in animals. Most practitioners (99%) felt that national guideline on rational antibiotic use is necessary and 87% believed a local antimicrobial guideline would be more useful than an international one. Around 80% disagreed with adding antibiotics with feed/water as a growth promoter in poultry and livestock. Regarding the major reasons of antibiotic resistance, irrational use of antibiotics was identified as the primary cause by 94.71%, followed by over-the-counter use, low dose, low-quality antibiotics, and waste disposal of antibiotics (Fig 1). Around 98% vets agreed that inappropriate or half course of antibiotics lead to antibiotic resistance. The lack of exposure and training regarding antibiotics and resistance had seen in middle aged vets. In addition, 48.2% poultry practitioners, 44.5% private veterinarians and 50.8% who had experienced from 1 to 3 years of practicing did not attend any training or workshop on AMR has impacts on attitude regarding antibiotics and resistance. Importantly, all aged group vets showed indifference attitude if individual or animal could not be treated with antibiotics whereas vets in different service (A5) and practicing (A5 and 6) showed significant attitude (Table 3).

**Table 3.**
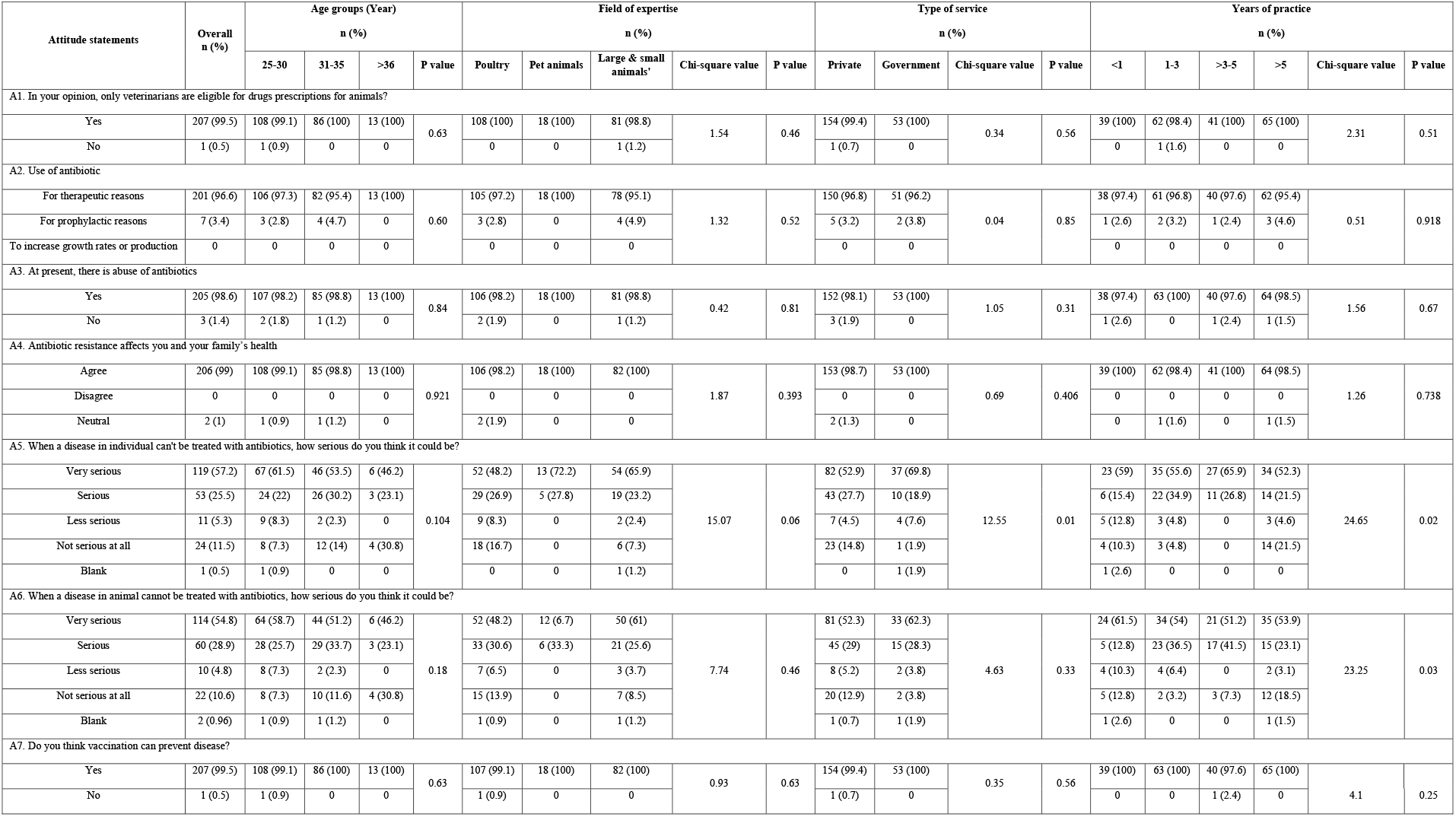
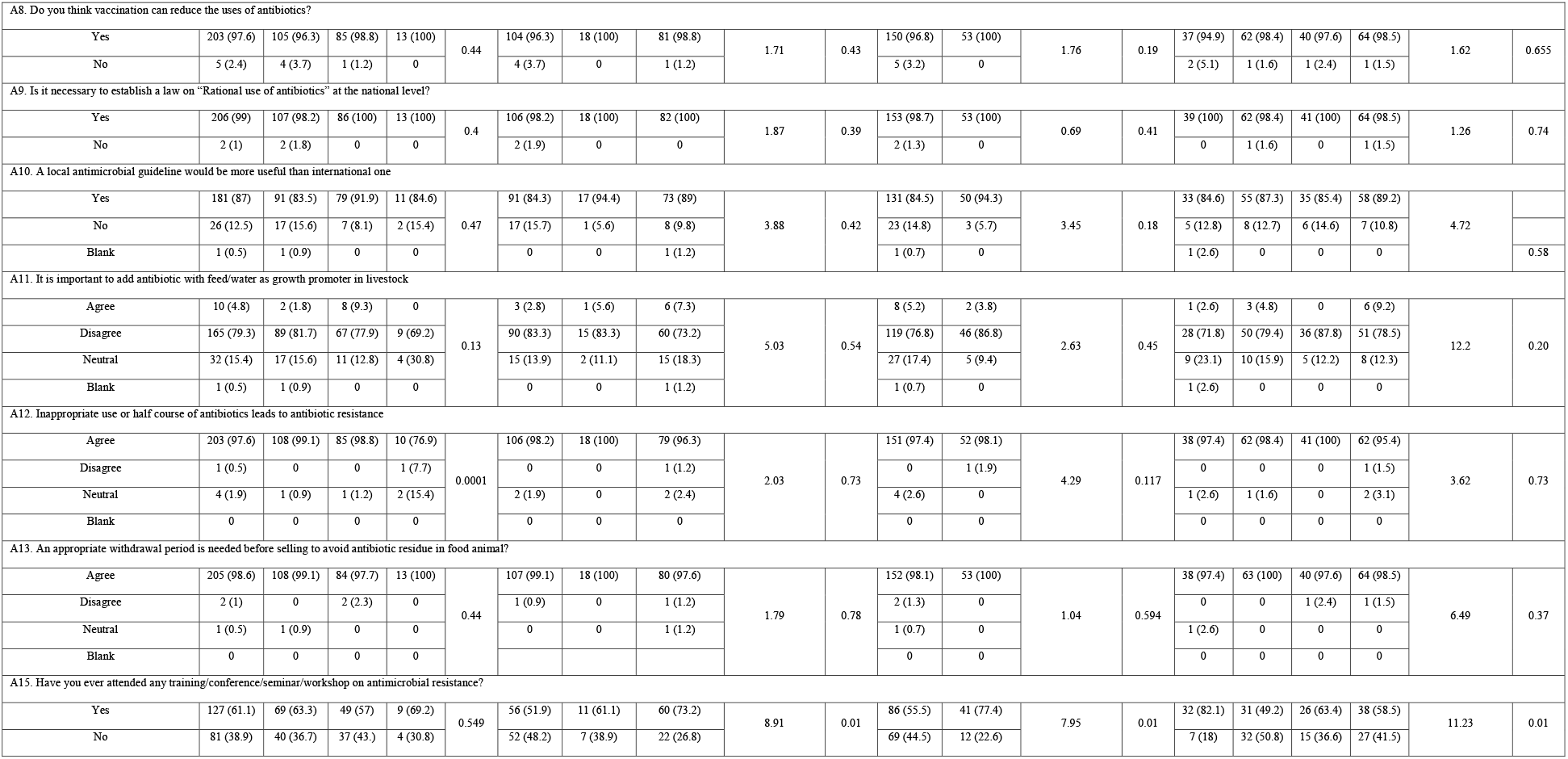
Practitioner’s attitude towards antibiotic use and resistance.

**Fig 1.** Major reasons of AMR indicated by the veterinarians

### The practice of the veterinarians regarding antibiotic prescribing

The majority (70.2%) of the veterinarians reported that they sometimes prescribe antibiotics over the phone or without examining the animals. Also, only 9.1% of the practitioners always or often recommend antimicrobial susceptibility testing before prescribing an antibiotic agents. Half of the participants prefer broad-spectrum antibiotics while the other half prefer narrow-spectrum antibiotics. Results also showed that antibiotics constitute a large percentage of daily prescribed drugs. Moreover, combined antibiotic therapy is also preferred to single therapy by about 65% of the practitioners, and old generation antibiotics are preferred to new generation antibiotics by most (63.5%) as a first-line treatment. Some practitioners (26%) reported prescribing antibiotics without determining the bodyweight of the animals. Most practitioners (74.5) do not mention the antibiotic withdrawal period in the prescriptions. When exploring the factors considered by the vets while prescribing antibiotics, the severity of the disease was found to be the most important factor. The vets also considered other factors such as availability of an antibiotic in the local market, culture sensitivity test report, economic status of the owner, and owners’ demand for antibiotics (Fig 2). There was significant variation in relation with practitioners age (P15 and 18), expertise (P4, 6, 11 and 13), type of service (P2, 10 and 17) and experienced (P2) and antibiotic practices (Table 4).

**Table 4.**
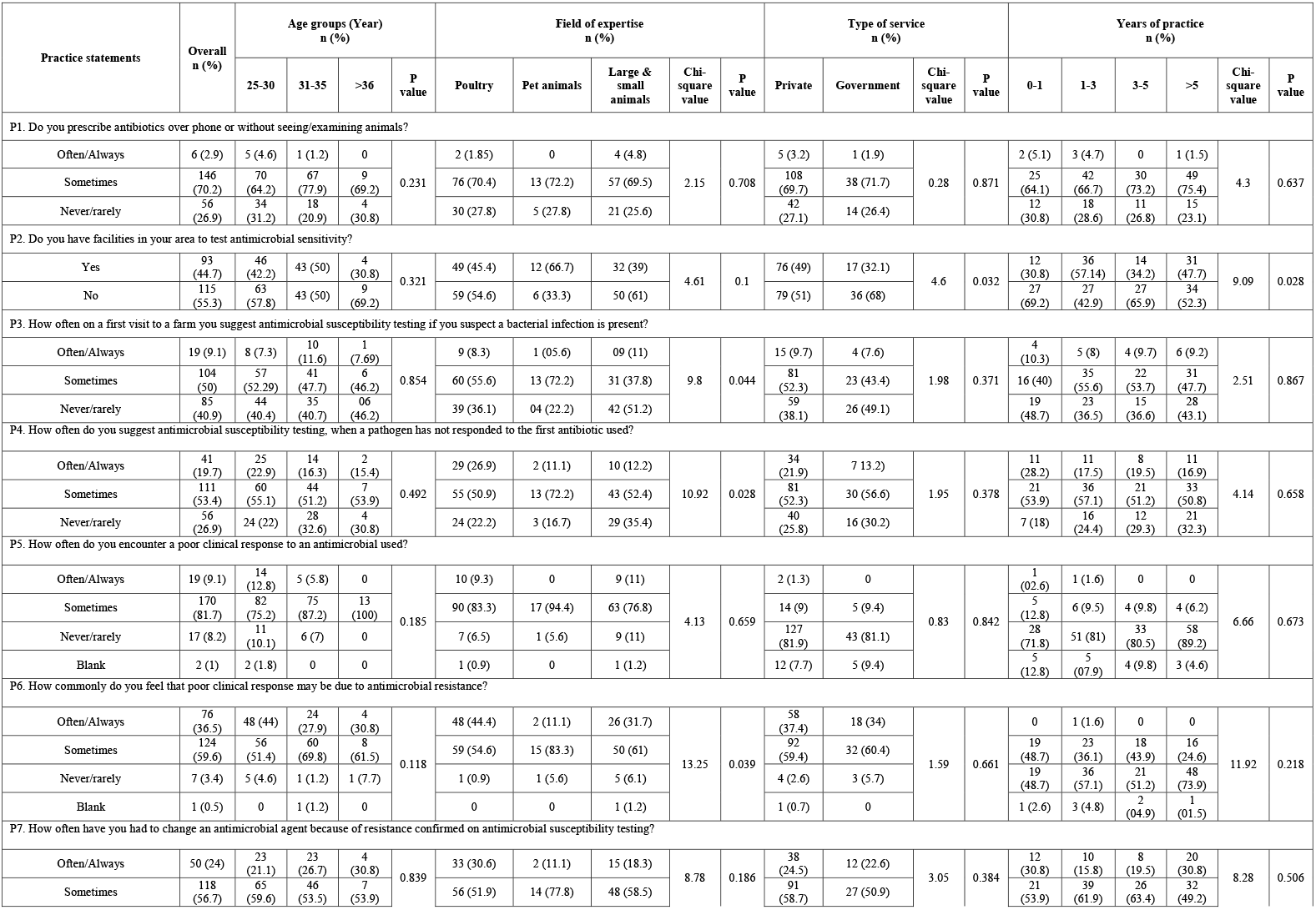
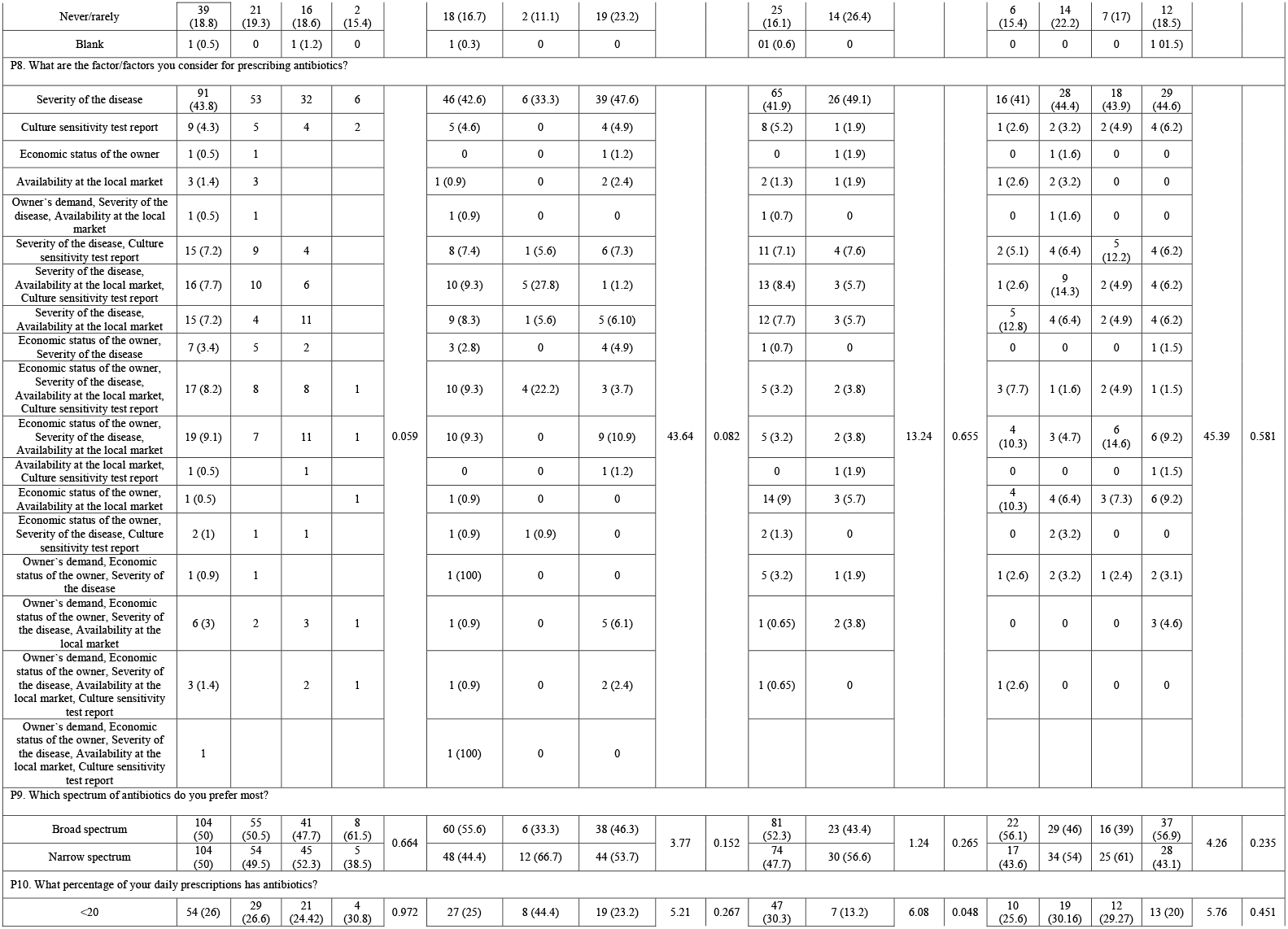
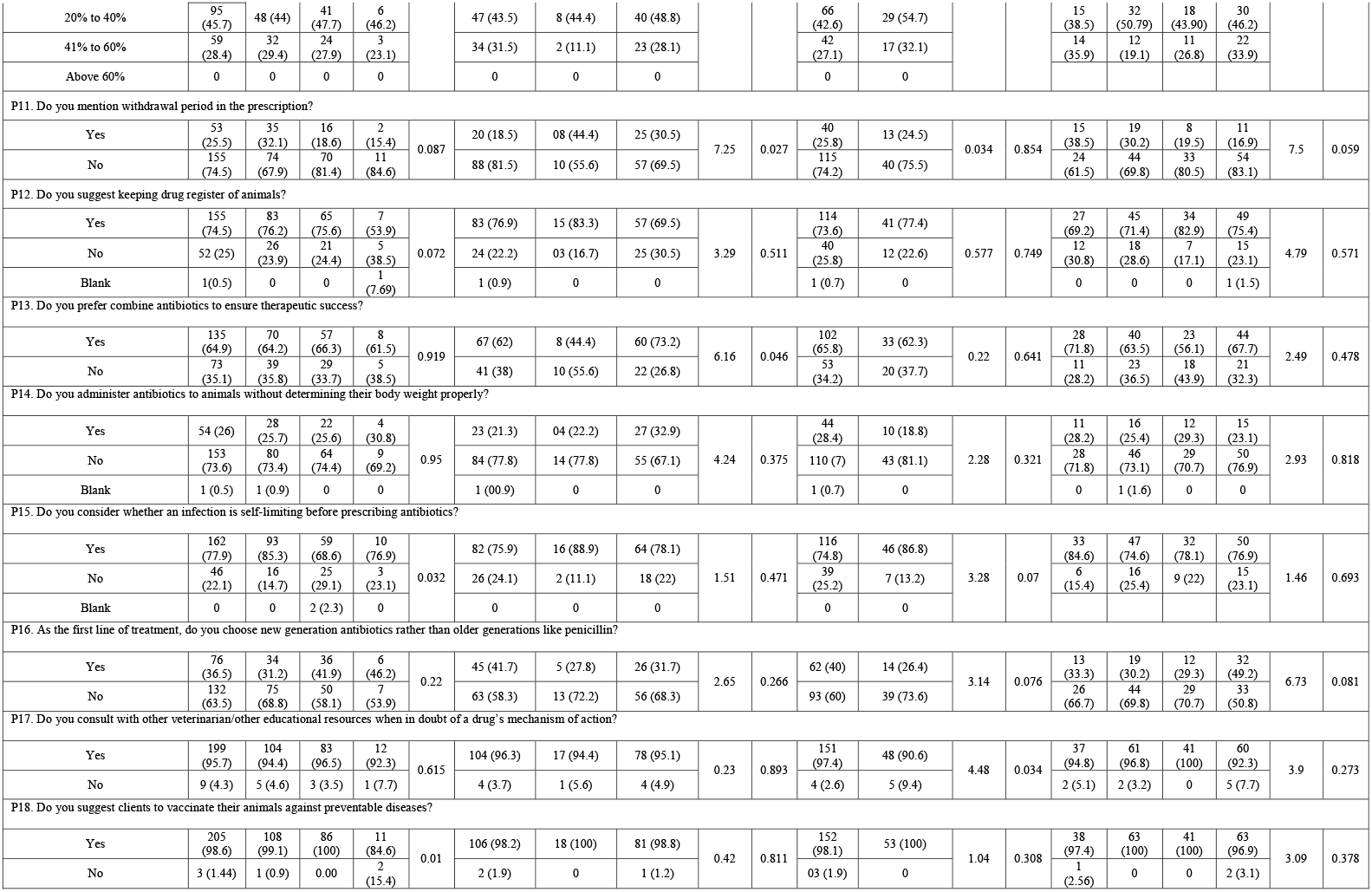
Practice of veterinary practitioner’s regarding antibiotic and resistance.

**Fig 2.** Factors considered by veterinarians while prescribing antibiotics

## Discussion

The present study explored the knowledge, attitude, and practice of the Bangladeshi veterinary practitioners regarding antibiotic use and resistance. It was found that some vets had gaps in knowledge in certain crucial concepts, for example, many practitioners (17.3%) considered antibiotics and antimicrobials to be the same. Failure to differentiate between antimicrobials and antibiotics and their roles can be a major reason for inappropriate antibiotic prescribing such as prescribing antibiotics for viral infections. Another surprising finding was that although most knew that antibiotics cannot cure viral infections, one-third of the vets believed antibiotics would speed up cold, cough, or other viral infections. However, there is no evidence that antibiotics can cure viral infections or speed up recovery of viral infections such as common cold [22,23].

Nearly all veterinarians were aware of antibiotic withdrawal period and considered it important to maintain an appropriate withdrawal period prior to selling animals treated with antibiotics in order to avoid antibiotic residues in animals. However paradoxically, while prescribing antibiotics, only one out of four practitioners mentioned the withdrawal period in the prescriptions. This may happen probably because practitioners do not have the knowledge of the withdrawal periods of the specific antibiotics they prescribe, or because they think the farmers will not understand or follow instructions related to withdrawal periods anyway. Besides, previous studies have shown that most Bangladeshi farmers do not have knowledge on antibiotic withdrawal period [10,11]. Hence, non adherence to the required withdrawal periods by either parties may result in the presence of residual antibiotics in food animal products [24]. Antibiotic residues can be toxic to humans as well as may contribute to the development of AMR [24,25].

Prescribing antibiotics based on the results of susceptibility testing is recommended to make sure that the prescribed regimen is effective against the infection. However, in this study, such practice was not often followed by the practitioners even after the initial treatment failed. This can partially be explained by the fact that most areas of Bangladesh did not have any facility to test antimicrobial sensitivity, as reported by the veterinarians. The absence of susceptibility data can also promote combined antibiotic therapy since the vets may want to prescribe more than one drug for maximizing the chance of therapeutic success with the hope that if one drug is found ineffective, the others will work. A study of the veterinary surgeons of the United Kingdom also reported similar findings where the surgeons only occasionally carried out susceptibility testing [26].

The use of antibiotics for disease prevention of animals by farmers and poultry dealers have been reported in Bangladesh [8], although such practices are not recommended [14]. Most veterinarians in this study do not consider the use of antimicrobials for disease prevention. Instead, the participants have shown a very positive attitude towards vaccination for both infection prevention and lowering the use of antibiotics. Given the fact that about half of the most significant animal diseases are of viral origins [27], vaccination can be very effective and efficient in lowering the occurrences of infectious diseases in animals and will subsequently confer financial gains to the farmers as well as help to minimize unnecessary use of antibiotics. Vaccines have also been recommended for infection prevention by WHO [14].

Participants were knowledgeable about antibiotic resistance, its causes, and its consequences. However, unless such knowledge is translated into practice, no real benefit will be achieved. We have identified a number of inappropriate practices by the veterinarians including excessive antibiotic prescribing, prescribing antibiotics over the phone without examining animals, not relying on susceptibility testing, not mentioning antibiotic withdrawal period in prescription, etc.

This survey revealed the varied difference in knowledge, attitude and practice of antibiotics use among different aged group of veterinarians in Bangladesh. It was not conclusively established the variation of predefined questions answer with the different aged group of vets. From this study it revealed that old aged vets with higher training or field based training have a higher knowledge of appropriate use of antibiotics and AMR. The difference has observed among vets aged groups that possibly could have the link to work experiences over time. To understand the perceptions and barriers, further investigation is required to appropriate use of antibiotics in livestock and in poultry among the vets subpopulation. It would help the policy makers and academicians to ensure proper training and impart practical field based knowledge of appropriate use of antibiotics and AMR to the vets students, young vets and all level aged groups of vets.

Another major problem is Bangladeshi farmers rely more upon village doctors, traditional healers and drug sellers and consider government veterinarians as the last resort for seeking health services for their livestock [9]. This trend needs to change and qualified veterinarians should be the primary source of advice in order to promote rational antibiotic use. Veterinarians should also play an active role in dispelling misconceptions of the farmers surrounding antibiotics, and themselves should adopt the appropriate practices. The government should focus on implementing the laws pertaining to judicious use of antibiotics, as well as recruiting more qualified veterinarians so that farmers can have easy access to them.

### Limitations

A few limitations were witnessed during conduct the current study. The number of participants in the survey was low that may be due to several factors such as unwillingness to participate or lack of internet access or long questionnaire or some questions seemed too difficult. Sometimes respondents may have declined to share information they considered inappropriate or mistaken, resulting in an under-reporting of certain aspects on antibiotics and AMR knowledge and practices. The study could not meet the exact proportional number of respondents with anticipated geographic locations due to freedom of choice of the respondents.

## Conclusions

The study findings suggest policy guidelines and advocacy to the public and private veterianrians in improving prudent use of antibiotics. Antimicrobial stewardship program in the public and private veterinary hospitals are needed to be initiated to promote the rational use of antibiotics. Improved knowledge and awareness of the veterinarians through continuous education and training can enhance the rational use of antibiotics. Dissemination of regularly updated national antibiotic use guidelines in food animals, understanding the role of good biosecurity and vaccination practices in disease prevention, including antimicrobial susceptibility testing at affordable costs with easy accessibility are the significant factors that need attention to combat the rising AMR in veterinary sector in Bangladesh.

## Data Availability

All relevant data are within the manuscript and its Supporting Information files

https://drive.google.com/file/d/12FWFhaFQHIxAaqfqOGOCSK-b6Uhwa_Q0/view?usp=sharing

## Supporting Information

**S1 Text. Questionnaire for KAP survey on antibiotics and AMR**.

## Acknowledgments

The author highly acknowledge the cooperation of the veterinarieans of Bangladesh. We also extend our gratitude to the lab members of ARAC, BLRI. We would like to thank Dr Md Ahaduzzaman for proofreading and copyediting our manuscript.

## Author Contributions

Conceptualization: Md Samun Sarker

Data curation: Md Samun Sarker, Ayesha Ahmed, Fatema Akter Mahua

Formal analysis: Iftekhar Ahmed, Shariful Islam

Investigation: Md Samun Sarker, Ruhena Begum

Methodology: Md Samun Sarker, Ruhena Begum

Software: Md Samun Sarker, Shariful Islam

Supervision: Mohammed A. Samad, Nure Alam Siddiky

Validation: Mohammed A. Samad, Nure Alam Siddiky

Visualization: Md Samun Sarker, Nure Alam Siddiky

Writing-original draft: Md Samun Sarker, Iftekhar Ahmed

Writing-review & editing: Mohammed A. Samad, Nure Alam Siddiky, Md Ehsanul Kabir, Ayesha Ahmed, Fatema Akter Mahua

## Disclosure of potential conflicts of interest

The authors declare that they have no conflict of interest.

## Notes

### Competing Interest Statement

The authors have declared no competing interest.

### Funding Statement

The author(s) received no specific
funding for this work

### Author Declarations

The study protocol was reviewed and approved by the Antimicrobial Resistance Action Center (ARAC), Animal Health Research Division, Bangladesh Livestock Research Institute (BLRI), Bangladesh (Approval no: 05/06/2020:06).

### Summary of Updates

The manuscript has been updated based on the reviewers comments

